# Transplantation and old stem cell age independently increase the risk of clonal hematopoiesis in long-term survivors of pediatric HCT

**DOI:** 10.1101/2024.09.28.24314531

**Authors:** K.F. Müskens, N. Wieringa, M. van Bergen, J.E. Bense, B.M. te Pas, A.P.J. de Pagter, A.C. Lankester, M.B. Bierings, D. Neuberg, S. Haitjema, L.C.M. Kremer, G.A. Huls, S. Nierkens, J.H. Jansen, C.A. Lindemans, A. de Graaf, M.E. Belderbos

**Affiliations:** Princess Máxima Center for Pediatric Oncology, Utrecht, The Netherlands; Laboratory of Hematology, Radboud University Medical Center, Nijmegen, The Netherlands; Division of Stem cell Transplantation, Department of Pediatrics, Willem-Alexander Children’s Hospital, Leiden University Medical Center, Leiden, Netherlands; Department of Data Science, Dana-Farber Cancer Institute, Boston, MA; Central Diagnostic Laboratory, University Medical Center Utrecht, Utrecht, The Netherlands; Department of Hematology, University Medical Center Groningen, Groningen, The Netherlands; Center for Translational Immunology, University Medical Center Utrecht, Utrecht, The Netherlands

## Abstract

In pediatric hematopoietic cell transplantation (HCT) recipients, transplanted donor cells may need to function far beyond normal human lifespan. Here, we investigated the risk of clonal hematopoiesis (CH) in 144 pediatric long-term HCT survivors, compared to 115 healthy controls. CH was detected in 16% of HCT survivors, at variant allele frequencies (VAFs) of 0.01-0.31. Mutations were predominantly in DNMT3A (80%) and TET2 (20%). Older stem cell age and the HCT procedure independently increased the risk of CH (odds ratios 1.07 per year increase (p<0.001) and 2.61 for HCT (p=0.02)), indicating both aging- and transplantation-induced effects. Large clones (VAF >0.10) were found exclusively in HCT recipients. Notably, CH was also detected within 15 years after cord blood HCT. Inflammatory processes around graft infusion were associated with CH presence. Future studies are required to track the evolution of post-transplant CH and its impact on future cardiovascular disease, second malignancies and overall survival.

**Significance statement:** As the number of long-term HCT survivors continues to increase, so does the population at risk of long-term effects. We demonstrate that pediatric HCT survivors are at increased risk of clonal hematopoiesis compared to the general population. Given the young age of these recipients, our data emphasize the need for prospective studies to assess the potential health consequences of post-transplant CH.

## Introduction

Clonal hematopoiesis (CH) refers to the detectable presence of a leukemia-associated mutation without overt hematologic malignancy^1–3^. In the general population, CH is strongly age-dependent and associated with an increased risk of developing myeloid malignancies, cardiovascular disease and overall mortality^1,2,4^. Previously, we showed that clonal expansion rates depend on the mutated gene^5^. Still, large variation exists in expansion rates, even in individuals with the same mutation^5,6^. Which mutation-independent processes drive the expansion of mutant clones remains incompletely understood.

Allogeneic hematopoietic cell transplantation (allo-HCT) provides a unique, controlled setting to study the impact of forced proliferation, inflammation and drug therapy on CH. Previous studies have shown that 1) hematopoietic stem cell (HSC) clones with CH driver mutations can be present in HCT donors, even at very young age^7,8^; 2) donor-derived HSCs with CH mutations can engraft in the recipient^7–11^; 3) clone size is generally increased in recipients compared to donors^9,11^; 4) the extent to which donor-derived mutant clones expand upon transplantation varies largely between individuals^7,8,10,11^; and 5) forced proliferation of HSCs during hematopoietic regeneration results in DNA damage and telomere shortening^10^, which may reduce long-term clonal diversity. While long-term effects are most relevant to HCT recipients transplanted at pediatric age given their long life-expectancy, most studies on post-transplant CH are limited to adults. Moreover, which transplant-related exposures facilitate the preferential outgrowth of CH-mutant over wildtype clones remains unstudied.

Here, using a well-annotated cohort of 144 long-term survivors of pediatric HCT, we investigated whether HCT at a young age increases the risk of developing CH and which extrinsic factors may confer a proliferative advantage to CH mutant clones.

## Methods

A total of 144 long-term survivors of allo-HCT (>5 years post-transplant), who underwent HCT during childhood (≤18 years) between 1981 and 2018, were enrolled during post-HCT follow-up at the Princess Máxima Center (PMC) from January 2022 to October 2023 (Supplementary Figure S1). Inclusion criteria were having received an allo-HCT at least five years prior. Exclusion criteria were inability to understand the patient informed consent forms. Patients had been transplanted at the UMC Utrecht/PMC (n=80) or Leiden University Medical Center (n=64). The study was approved by the local ethics committee (NedMec no. NL77721.041.21) in accordance with the declaration of Helsinki. All patients or their caretakers provided written informed consent. Clinical characteristics and demographics were extracted from medical charts. CH was assessed in whole blood collected at study enrolment using a panel targeting 27 myeloid and lymphoid malignancy driver genes, as previously described^5^. Somatic variants were called at variant allele frequency (VAF) ≥0.01 and ≥10 mutant unique reads. To prevent detection of germline variants, variants at VAF >0.40 were filtered. Post-transplant CH was compared to CH within leftover graft material from healthy donors, unrelated to our survivor cohort^12^. Both cohorts were processed and analyzed using the same pipelines. Associations between clinical characteristics and CH were tested using Wilcoxon, Chi-squared or Fisher’s exact tests as appropriate. Logistic and linear regression models were used to identify risk factors for the prevalence of CH and VAF, respectively, correcting for potential confounders. Post-transplant viral reactivations were defined as cytomegalovirus, Epstein-Barr-virus and/or adenovirus loads >1000 copies/mL during pre-emptive screening. For patients transplanted in the UMC Utrecht/PMC, historical laboratory data, including weekly pre-emptive C-reactive protein (CRP) measurements around graft infusion, were available and extracted via the Utrecht Patient Oriented Database^13^.

## Results

### CH prevalence is higher in long-term survivors of pediatric HCT compared to healthy controls

A total of 144 HCT recipients were enrolled, at a median calendar age of 20.5 years (interquartile range (IQR): 16.2-27.7, Table 1). HSC age (median: 32.1 years, IQR 15.6-43.7) was calculated by summing the donor age at HCT (median: 13.5 years, IQR: 5.1-26.8) and the time between HCT and CH assessment (“follow-up time”, median: 10.4 years, IQR: 8-17.9). In total, we identified 30 CH mutations with VAF ≥0.01 in 23 HCT recipients (16%; 95% confidence interval (CI): 10-23%, Figure 1B). Mutations were exclusively found in *DNMT3A* (24 mutations, 80%) or *TET2* (6 mutations, 20%). Most were single-base substitutions (23 mutations, 77%), of which 12 were cytosine-to-thymine transitions (Supplementary Figure S2). VAFs ranged from 0.01 to 0.31, with a median of 0.02. Chimerism analysis confirmed that post-transplant CH originated from donor cells in 20 out of 23 cases. In the remaining recipients, recipient origin of CH could not be excluded due to residual patient chimerism exceeding the size of the CH clone.

**Table 1.** HCT characteristics for study cohort. P values calculated from Chi-squared test or Fisher-exact test if expected count <5 (categorical variables) or Wilcoxon rank-sum test (continuous variables). Serotherapy consisted of either anti-thymocyte globulin or alemtuzumab (Campath). Abbreviations: CH: clonal hematopoiesis; IQR: interquartile range; HCT: Hematopoietic stem cell transplantation; HSC: hematopoietic stem cell; TBI: Total body irradiation.

| Determinant | Total<br>(n = 144) |  | CH<br>(n = 23) |  | No CH<br>(n = 121) |  | P<br>value |
| --- | --- | --- | --- | --- | --- | --- | --- |
| **Recipient sex** |  |  |  |  |  |  |  |
| Male | 83 | (58%) | 13 | (57%) | 70 | (58%) | 1 |
| **Recipient age at Inclusion;** (years, median, IQR) | 20.5 | 16.2-27.7 | 28.3 | (19.4-33.2) | 20.2 | (15.9-25.7) | 0.03 |
| **Recipient age at HCT;** (years, median, IQR) | 8.0 | 4.0-12.3 | 7.3 | (3.6-15.3) | 8.0 | (4.1-6.7) | 0.59 |
| **HSC age;** (years, median, IQR) | 32.1 | 15.6-43.7 | 44.0 | (34.7-51.4) | 29.6 | (14.2-42.0) | <0.01 |
| **Donor age at HCT;** (years, median, IQR) | 13.5 | 5.1-26.8 | 30.2 | (19.6-36.4) | 11.6 | (4.3-25.1) | <0.01 |
| **Follow-up time;** (years, median, IQR) | 12.1 | 8.0-17.9 | 18.4 | (8.5-22.8) | 11.6 | (7.9-16.1) | 0.07 |
| **Underlying disease** |  |  |  |  |  |  | 0.33 |
| Hemato-oncology | 116 | (81%) | 21 | (91%) | 95 | (79%) |  |
| Bone marrow failure syndromes | 24 | (17%) | 2 | (9%) | 22 | (18%) |  |
| Immunodeficiency | 4 | (3%) | 0 | (0%) | 4 | (3%) |  |
| **Stem cell source** |  |  |  |  |  |  | 0.01 |
| Bone marrow | 101 | (70%) | 18 | (78%) | 83 | (69%) |  |
| Cord blood | 33 | (23%) | 1 | (4%) | 32 | (26%) |  |
| Peripheral blood | 10 | (7%) | 4 | (17%) | 6 | (5%) |  |
| **Relation to donor** |  |  |  |  |  |  | 0.21 |
| Unrelated | 84 | (58%) | 16 | (70%) | 68 | (56%) |  |
| Family | 60 | (42%) | 7 | (30%) | 53 | (44%) |  |
| Sibling | 53 | (37%) | 5 | (22%) | 48 | (40%) |  |
| Parent | 7 | (5%) | 2 | (9%) | 5 | (4%) |  |
| **Conditioning** |  |  |  |  |  |  |  |
| Chemotherapy-based | 112 | (78%) | 18 | (78%) | 99 | (78%) | 1 |
| TBI-based | 32 | (22%) | 5 | (22%) | 27 | (22%) |  |
| Myeloablative | 134 | (93%) | 20 | (87%) | 114 | (94%) | 1 |
| Non-myeloablative | 10 | (7%) | 3 | (13%) | 7 | (6%) |  |
| **Serotherapy** |  |  |  |  |  |  | 0.01 |
| Yes | 87 | (60%) | 20 | (87%) | 68 | (56%) |  |
| No | 54 | (38%) | 3 | (13%) | 53 | (44%) |  |

**Figure 1.**
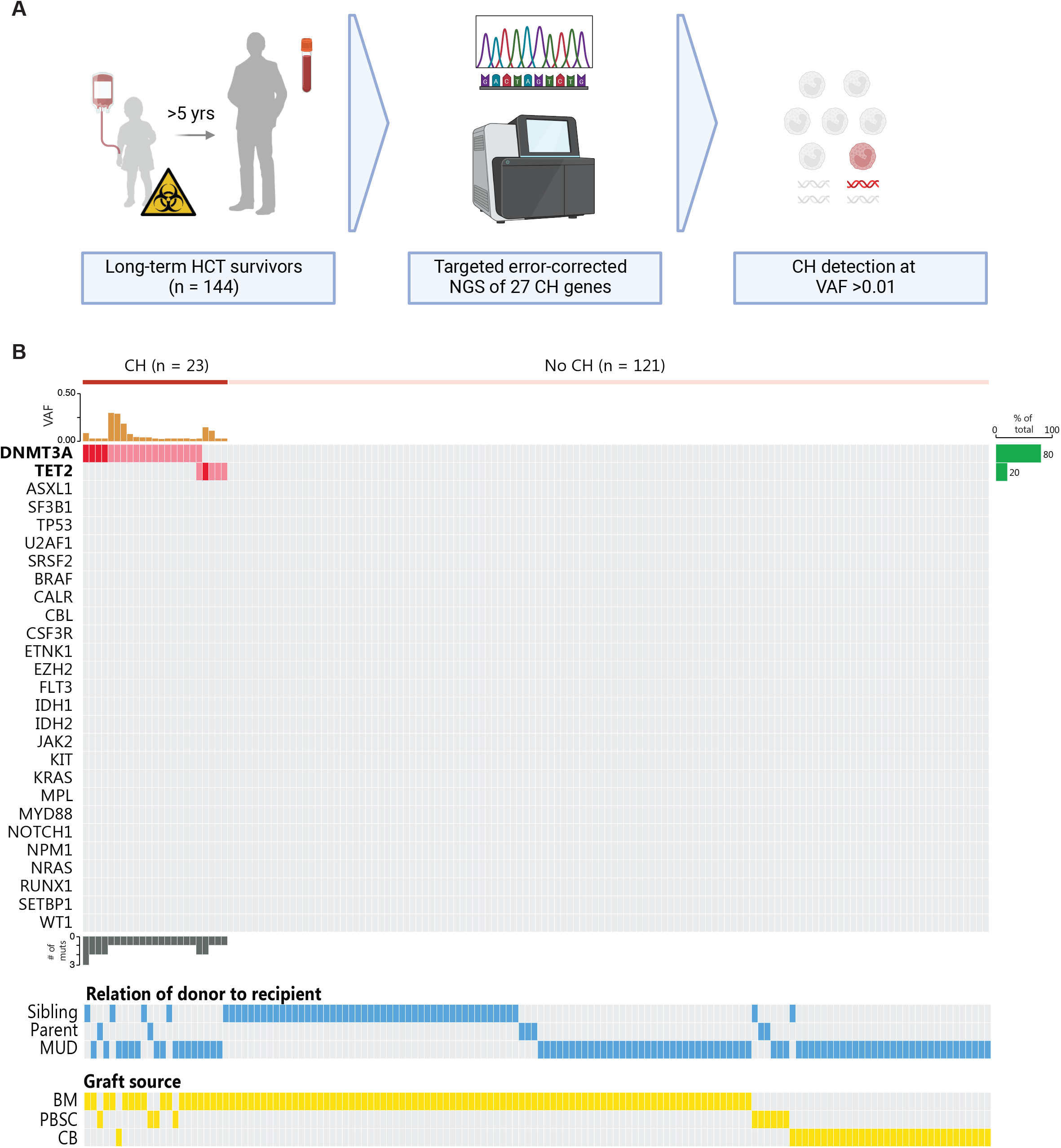
CH is common after HCT at pediatric age. **A)** Study design. **B)** Oncoprint of all HCT recipients, sorted by identified CH mutation (red) and variant allele frequency (VAF, orange bars). A darker shade indicates multiple mutations in the same gene. Relation to donor (blue) and stem cell source (yellow) are indicated. *Abbreviations: HCT: hematopoietic cell transplantation; NGS: next generation sequencing; CH: clonal hematopoiesis; VAF: variant allele frequency; MUD: matched unrelated donor; BM: bone marrow; PBSC: peripheral blood stem cells; CB: cord blood*.

To determine whether HCT affects the prevalence and characteristics of CH, we compared our HCT cohort to 115 healthy, untransplanted controls (median age: 48.0 years, IQR: 34.0-56.5, Supplementary Table S1, Supplementary Figure S3). In line with previous studies^1,2,14,15^, CH was virtually absent in healthy individuals <40 years. In contrast, eight HCT recipients with HSC-ages <40 years had detectable CH (Figure 2A). In both cohorts, older HSC age was associated with increased risk of CH (odds ratio 1.07 per year increase, p<0.001, Figure 2A-B and Supplementary Figure S3D). When comparing individuals with similar HSC ages, the age-corrected prevalence of CH was significantly higher in HCT recipients compared to controls (Figure 2A). In the HCT setting, HSC age consists of donor age, reflecting the risk of CH-mutant HSCs being present in the graft, and follow-up, reflecting the time during which mutant HSCs expand or arise in the recipient (Figure 2C-D). Additionally, the calendar age of the recipient at HCT represents the age of the niche receiving the donor HSCs (Figure 2E). In multivariable regression, both donor age and follow-up time were significant predictors of CH (Figure 2F), whilst recipient age at HCT was not (p=0.68).

**Figure 2.**
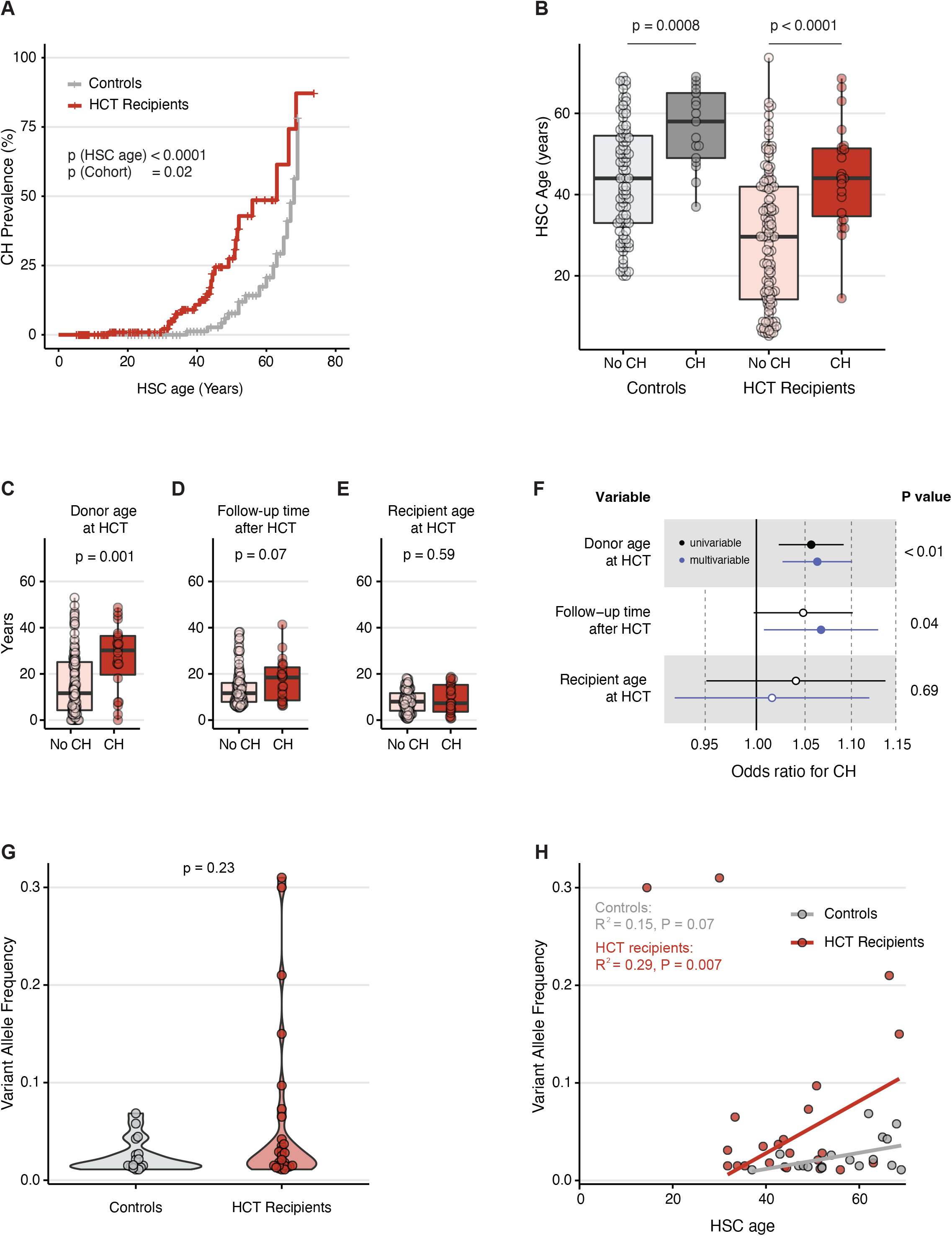
Comparison of CH in HCT recipients and controls. **A)** Cumulative prevalence of CH plotted against HSC age, split by cohort. P values are derived from a multivariable logistic regression model with cohort (HCT recipient versus control) and HSC age as determinants and CH status as outcome. **B)** Bar plot of HSC age in individuals with and without CH for both cohorts. **C-E)** Bar plots of donor age, follow-up time and recipient age in HCT recipients with and without CH. **F)** Forest plot depicting the effect of timespans from C-E on CH prevalence in logistic regression models with CH status as outcome. The multivariate model includes all three timespans. **G)** Violin plot of variant allele frequencies (VAF) of CH clones in controls and HCT recipients. **H)** Scatterplot of VAF of largest CH clone per individual versus HSC age. Linear regression is calculated for each cohort separately, excluding the outliers with VAF >0.30 in the HCT Recipient cohort. P values in B-E and G are calculated using Wilcoxon rank-sum test. *Abbreviations: HCT: hematopoietic cell transplantation; HSC: hematopoietic stem cell; CH: clonal hematopoiesis*.

Most CH clones were small, both in HCT recipients and controls (median VAF 0.023 and 0.015 respectively, p=0.23, Figure 2G). Notably, very large clones with VAF >0.10 were exclusively detected in HCT recipients. Although a trend of increased clone size at higher HSC age was observed for both HCT recipients and controls (Figure 2H), this only explained part of the observed variation in clone sizes. In particular, two HCT recipients carried DNMT3A-mutated clones with VAFs of 0.30 and 0.31, at HSC ages of 14.5 and 30.0 years, respectively. Strikingly, the former of these two recipients was transplanted with cord blood, suggesting that even cord blood grafts may contain HSCs with CH driver mutations. Collectively, these data demonstrate that HCT recipients are at increased risk of CH at younger HSC age compared to controls, likely originating from mutant donor HSPCs which preferentially expand in the context of HCT and persist into adulthood.

### Systemic inflammation is associated with post-transplant CH

To identify HCT-related exposures that may contribute to the development of CH, we compared clinical characteristics between HCT recipients with and without CH. No differences were observed in underlying disease, stem cell source, CD34 dose or donor-recipient relation (Figure 3A). Remarkably, we identified various post-transplant exposures associated with presence of CH. First, CH prevalence was higher in recipients who had received serotherapy as part of their conditioning regimen. After serotherapy administration (usually day -9 to -5 before graft infusion), T-cell lysis can result in an inflammatory response marked by fever and increased C-reactive protein (CRP)^16^. To investigate whether HCT recipients with long-term CH had experienced such an inflammatory response, we investigated CRP values around time of graft infusion. Hereto, we extracted CRP values, obtained during pre-emptive monitoring, for all HCT recipients transplanted at the UMC Utrecht/PMC (n=80, Figure 3B-C, Supplementary Figure S4). Within this subset, all HCT recipients that later developed CH had received serotherapy. As expected, serotherapy was associated with a transient CRP rise (Supplementary Figure S4B). Strikingly, CRP levels before and shortly after graft infusion were higher for HCT recipients with subsequent CH compared to those without, both in the overall cohort and within those who had received serotherapy. This difference was not observed at later timepoints. In addition, post-transplant viral reactivations, defined as cytomegalovirus, Epstein-Barr-virus and/or adenovirus loads >1000 copies/mL during pre-emptive screening, were associated with CH in univariable analysis (Figure 3D). Whether this is the consequence of serotherapy or an independent inflammatory event could not be established. We did not observe a significant association between acute graft-versus-host disease (aGvHD) and CH (Figure 3E). None of the HCT recipients in our cohort had developed a secondary hematologic malignancy at time of enrollment.

**Figure 3.**
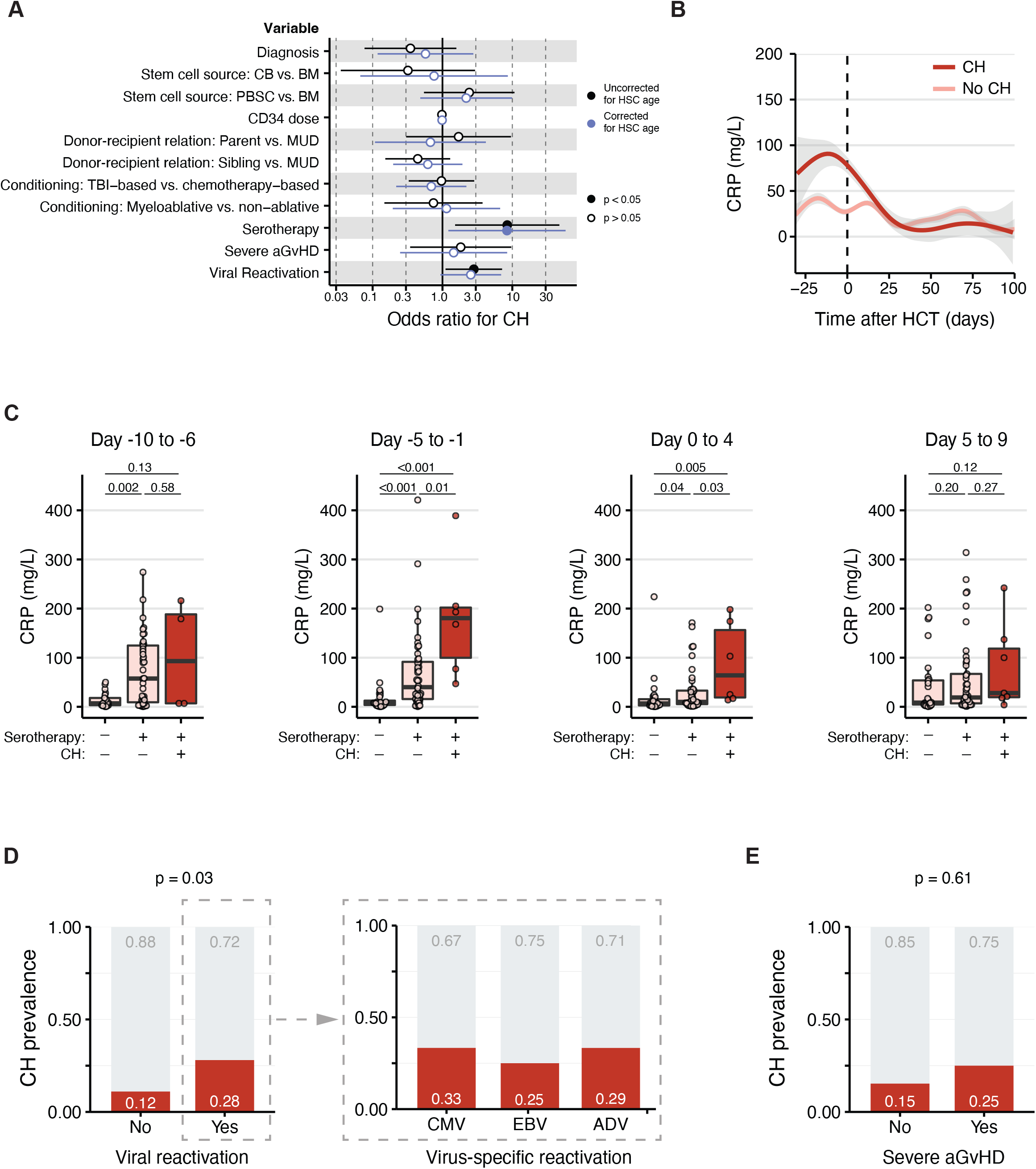
Clinical factors associated with prevalence of CH. **A)** Forest plot showing the effect of clinical characteristics on CH prevalence of univariable and multivariable logistic regression models. Stem cell source was corrected for donor age, CD34 dose was corrected for stem cell source and serotherapy was corrected for relation to donor. **B)** CRP measurements around HCT in individuals with and without CH. Lines indicate LOESS regression with 95% confidence interval. **C)** Box plots demonstrating the highest CRP measured within 5-day intervals around graft infusion, split by serotherapy (yes/no) and CH status. P values are calculated using Wilcoxon rank-sum test. **D-E)** Prevalence of CH in individuals with and without viral reactivations, including CMV, EBV and adenovirus reactivation (D) and with and without severe acute graft-versus-host disease (E). P values are calculated using Chi-squared test. *Abbreviations: CH: clonal hematopoiesis; CB: cord blood; BM: bone marrow; PBSC: peripheral blood stem cells; MUD: matched unrelated donor; TBI: total body irradiation; aGvHD: acute graft-versus host-disease; HSC: hematopoietic stem cell; CRP: C-reactive protein; HCT: hematopoietic cell transplantation; CMV: cytomegalovirus; EBV: Epstein-Barr virus; ADV: adenovirus*.

## Discussion

Due to the increasing use of HCT as a treatment and improvements in supportive care protocols, the number of long-term HCT survivors continues to increase. While much research has focused on the non-hematopoietic late effects after HCT^17^, the long-term effects on the hematopoietic system – the only organ not directly exposed to chemotherapy – remain unclear. Long-term complications are particularly relevant for pediatric recipients, who have a life expectancy of several decades after successful HCT. Here, we demonstrate that long-term survivors of pediatric HCT are at increased risk of CH compared to the general population. Importantly, older HSC age and the HCT procedure itself were independently associated with post-transplant CH, suggesting the involvement of both aging- and transplantation-induced effects. Furthermore, consistent with existing pre-clinical data indicating that inflammation promotes clonal expansion^18^, we found an association between post-transplant CH and (serotherapy-induced) inflammation around graft infusion. This supports the hypothesis that systemic inflammation might promote the selection and/or preferential expansion of mutant HSCs, resulting in detectable CH years later.

Our findings are in line with previous studies in adult HCT cohorts, demonstrating that the prevalence of CH is higher in HCT recipients compared to their donors^19^. Using sensitive backtracking studies, most cases of adult post-transplant CH could be traced back to the donor graft, even in very young donors^7^. Furthermore, donor-derived CH clones were more likely to engraft and/or expand after HCT, resulting in an increased clone size in recipients compared to donors^9,11^. While residual graft material was not available for our study cohort, these transplant donors would be very unlikely to harbor detectable CH clones given their young age. It is important to note that the detection of CH is highly dependent on the sequencing thresholds used, and that CH-negative individuals may still harbor small CH clones below the lower limit of detection. Accordingly, the absence of detectable CH in the HCT donor does not rule out the possibility of mutated HSCs already being present in the graft and becoming detectable in the recipient after HCT. In line with this notion, we found a positive association between donor age and the risk of post-transplant CH. Notably, we also observed CH within 15 years after UCB HCT, which is amongst the youngest cases of CH reported thus far. This is consistent with previous single-cell lineage-tracing^20^ and twin studies^21^, suggesting that HSCs carrying leukemia-associated mutations might already be present at birth, and only grow out to become detectable clones upon HCT, which creates an environment favoring the selective expansion of mutant HSCs.

The clinical impact of post-transplant CH remains to a large part unknown. Long-term HCT survivors are confronted with various potential late effects, including second malignancies and cardiovascular disease^22,23^, which may be affected by or related to the presence of CH^24,25^. In our cohort, we did not observe any individuals with donor-derived myelodysplasia or secondary hematologic malignancies. Yet, the detection of these complications is better addressed in prospective rather that cross-sectional studies. Notably, we have recently shown that the risk of malignant transformation of CH depends on the mutated gene as well as its growth dynamics^5^. While the observed prevalence of post-transplant CH in young HSCs indicates either a transient or persistent increase in clonal growth, discriminating between these two may identify clones with different risk of malignant progression. Future research should focus on longitudinal monitoring of CH in allo-HCT recipients to investigate growth dynamics and better assess the long-term clinical consequences, including cardiovascular disease, second malignancies and overall survival.

Our study has several limitations, including heterogeneity in the patient population and time of sampling, and the unavailability of pre-transplant graft material. Additionally, by focusing on very long-term survivors, HCT recipients with severe post-HCT complications may be underrepresented in our study cohort, as these complications may impact their long-term survival and/or ability to enroll. Accordingly, the lack of association between CH and aGvHD needs to be interpreted with caution. Yet, from a clinical perspective, our findings remain relevant, since long-term HCT related effects are only relevant for those who survive. Finally, the association between CH, serotherapy, CRP and post-HCT viral reactivations does not indicate causality. In fact, many of these processes are linked, for example, serotherapy-induced T-cell depletion is associated with increased risk of viral reactivations. In addition, the presence of CH itself may affect the HCT-associated inflammatory processes. Future studies, including longitudinal sampling of both donor and recipient, may help understand how different aspects of HCT (e.g., mobilization, engraftment, forced proliferation, inflammation) affect the prevalence and dynamics of post-transplant CH.

In summary, pediatric HCT recipients are at increased risk of CH at a young age, emphasizing the importance of prospective screening. Insight into the longitudinal dynamics of CH post-HCT will be crucial to understanding the underlying mechanisms and to determine its potential impact on post-transplant health.

## Data Availability

All data produced in the present study are available upon reasonable request to the authors

## Acknowledgements

This study was financially supported by a VENI grant of the Netherlands Organization for Scientific Research (NWO) (grant number VI.Veni.202.021 to MEB), a physician scientist grant of the European Hematology Association (to MEB), a John Hansen research grant from the DKMS and a KiKa project grant (project number 418). The KiKa project grant supported the salary of KFM. The funders had no role in the design, execution or writing of this work, nor in the decision to submit results. We would like to thank M.C.H. de Groot for extracting historical laboratory data from the Utrecht Patient Oriented Database.

## Authorship Contributions

Conceptualization: All authors. Data analysis and figures: KFM, MvB, AdG, MEB. Writing (original draft): KFM and MEB. Writing (review and editing): All authors. All authors have read and agreed to the published version of the manuscript.

## Reference list

1. Jaiswal, S. et al. Age-Related Clonal Hematopoiesis Associated with Adverse Outcomes. N. Engl. J. Med. 371, 2488–2498 (2014).

2. Genovese, G. et al. Clonal Hematopoiesis and Blood-Cancer Risk Inferred from Blood DNA Sequence. N. Engl. J. Med. 371, 2477–2487 (2014).

3. Jaiswal, S. & Ebert, B. L. Clonal hematopoiesis in human aging and disease. Science (80-.). 366, (2019).

4. Zink, F. et al. Clonal hematopoiesis, with and without candidate driver mutations, is common in the elderly. Blood 130, 742–752 (2017).

5. van Zeventer, I. A. et al. Evolutionary landscape of clonal hematopoiesis in 3,359 individuals from the general population. Cancer Cell 41, 1017-1031.e4 (2023).

6. Fabre, M. A. et al. The longitudinal dynamics and natural history of clonal haematopoiesis. Nat. 2022 6067913 606, 335–342 (2022).

7. Wong, W. H., Bhatt, S. & Trinkaus, K. Engraftment of rare, pathogenic donor hematopoietic mutations in unrelated hematopoietic stem cell transplantation. Sci. Transl. Med. 12, (2020).

8. Gibson, C. J. et al. Donor Clonal Hematopoiesis and Recipient Outcomes After Transplantation. J. Clin. Oncol. 40, 189 (2022).

9. Boettcher, S. et al. Clonal hematopoiesis in donors and long-term survivors of related allogeneic hematopoietic stem cell transplantation. Blood 135, 1548–1559 (2020).

10. Campbell, P., Chapman, M. S., Wilk, M. & Boettcher, S. Clonal dynamics after allogeneic haematopoietic cell transplantation using genome-wide somatic mutations. doi:10.21203/rs.3.rs-2868644/v1.

11. Frick, M. et al. Role of donor clonal hematopoiesis in allogeneic hematopoietic stem-cell transplantation. J. Clin. Oncol. 37, 375–385 (2019).

12. van Bergen, M. G. J. M. et al. Characterization of a genomic region 8 kb downstream of GFI1B associated with myeloproliferative neoplasms. Biochim. Biophys. Acta - Mol. Basis Dis. 1867, 166259 (2021).

13. Ten Berg, M. J. et al. Linking laboratory and medication data: New opportunities for pharmacoepidemiological research. Clin. Chem. Lab. Med. 45, 13–19 (2007).

14. Xie, M. et al. Age-related cancer mutations associated with clonal hematopoietic expansion. Nat. Med. 20, 1472 (2014).

15. Abelson, S. et al. Prediction of acute myeloid leukaemia risk in healthy individuals. Nature 559, 400–404 (2018).

16. Brodska, H. et al. Marked increase of procalcitonin after the administration of anti-thymocyte globulin in patients before hematopoietic stem cell transplantation does not indicate sepsis: a prospective study. Crit. Care 13, R37–R37 (2009).

17. Baker, K. S., Bresters, D. & Sande, J. E. The burden of cure: Long-term side effects following Hematopoietic Stem Cell Transplantation (HSCT) in children. Pediatric Clinics of North America vol. 57 323–342 at 10.1016/j.pcl.2009.11.008 (2010).

18. Florez, M. A. et al. Clonal hematopoiesis: Mutation-specific adaptation to environmental change. Cell Stem Cell 29, 882–904 (2022).

19. Kim, K. H. et al. Clonal hematopoiesis in the donor does not adversely affect long-term outcomes following allogeneic hematopoietic stem cell transplantation: result from a 13-year follow-up. Haematologica 108, 1817–1826 (2023).

20. Williams, N. et al. Life histories of myeloproliferative neoplasms inferred from phylogenies. Nature 602, 162–168 (2022).

21. Ford, A. M., Colman, S. & Greaves, M. Covert pre-leukaemic clones in healthy co-twins of patients with childhood acute lymphoblastic leukaemia. Leuk. 2022 371 37, 47–52 (2022).

22. Carreras, E., Dufour, C., Mohty, M. & Kröger, N. The EBMT Handbook: Hematopoietic Stem Cell Transplantation and Cellular Therapies [Internet]. EBMT Handb. Hematop. Stem Cell Transplant. Cell. Ther. 1–702 (2019) doi:10.1007/978-3-030-02278-5.

23. Armenian, S. H. & Chow, E. J. Cardiovascular disease in survivors of hematopoietic cell transplantation. Cancer 120, 469 (2014).

24. Rhee, J. W. et al. Clonal Hematopoiesis and Cardiovascular Disease in Patients with Multiple Myeloma Undergoing Hematopoietic Cell Transplant. JAMA Cardiol. 9, 16–24 (2024).

25. Kato, M. et al. Donor cell-derived hematological malignancy: A survey by the Japan Society for Hematopoietic Cell Transplantation. Leukemia 30, 1742–1745 (2016).

