## Supplementary files for "Transplantation and old stem cell age independently increase the risk of clonal hematopoiesis in long-term survivors of pediatric HCT"

Supplementary Table S1

| Determinant | Total (n = 115) | | CH (n = 17) | | No CH (n = 98) | | P value |
|---|---|---|---|---|---|---|---|
| **Recipient sex** | | | | | | | |
| Male | 76 | (66%) | 9 | (53%) | 66 | (67%) | 0.38 |
| **Age;** (years, median, IQR) | 48.0 | (34.0-56.5) | 58.0 | (49.0-65.0) | 44.0 | (33.0-54.8) | <0.001 |

***Supplementary table S1. Demographics of control cohort.*** *P values calculated from Chi-squared test (categorical variables) or Wilcoxon rank-sum test (continuous variables). Abbreviations: CH: clonal hematopoiesis; IQR: interquartile range.*

Supplementary Figure S1

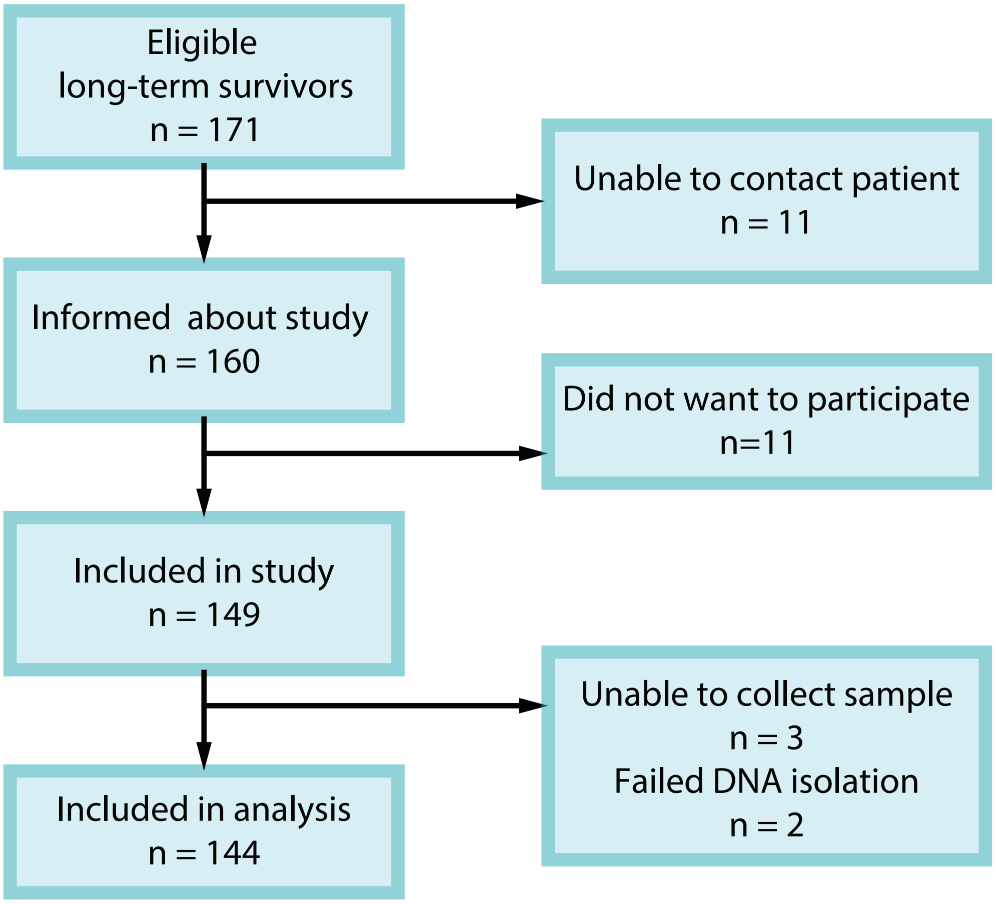


Overview of screening and study inclusion.

Supplementary Figure S2

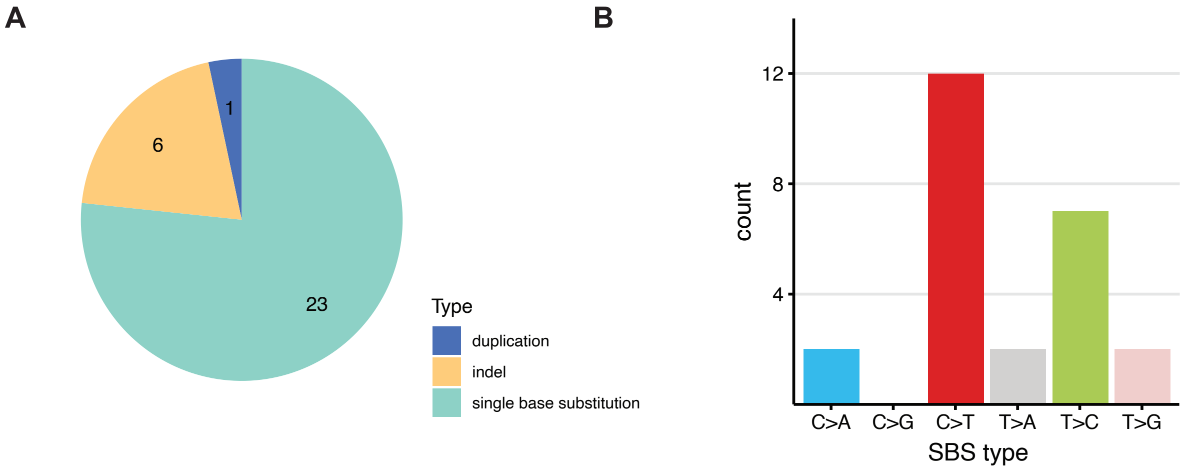


A) Pie chart depicting the type of mutations for all CH mutations within the HCT recipient cohort.
B) Type of single base substitution (SBS) for SBS mutations within the HCT recipient cohort.

*Abbreviations: SBS: single base substitution.*

Supplementary Figure S3

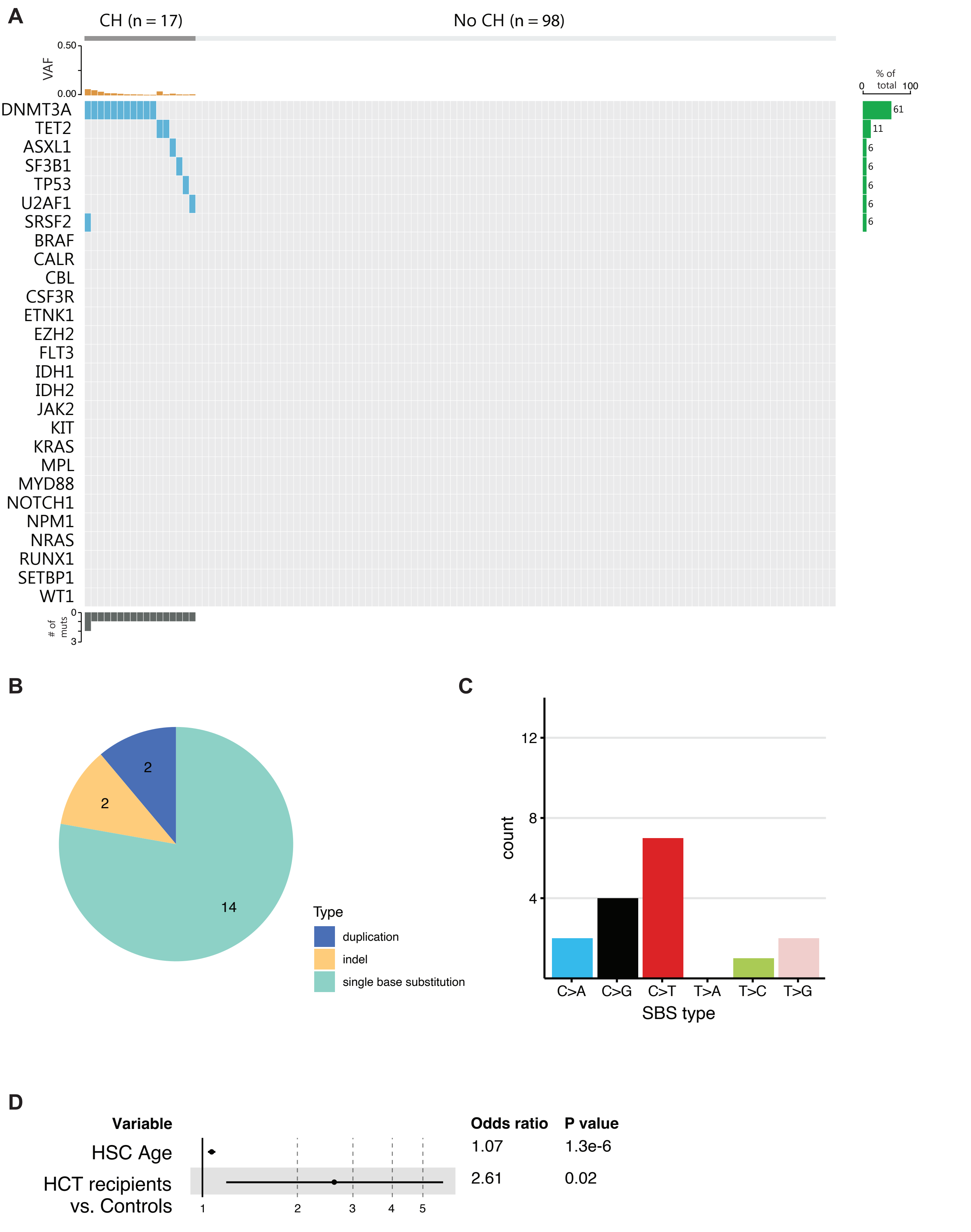


A) Oncoprint of all healthy controls, sorted by identified CH mutation (blue) and variant allele frequency (VAF, orange bars).

B) Pie chart depicting the type of mutations for all CH mutations within the control cohort.

C) Type of single base substitution (SBS) for SBS mutations within the control cohort.

D) Forest plot showing results from multivariable logistic regression model, with HSC age and cohort (HCT recipients versus control) as determinants and CH status at outcome.

*Abbreviations: CH: clonal hematopoiesis; VAF: variant allele frequency; SBS: single base substitution; HSC: hematopoietic stem cell; HCT: hematopoietic cell transplantation.*

Supplementary Figure S4

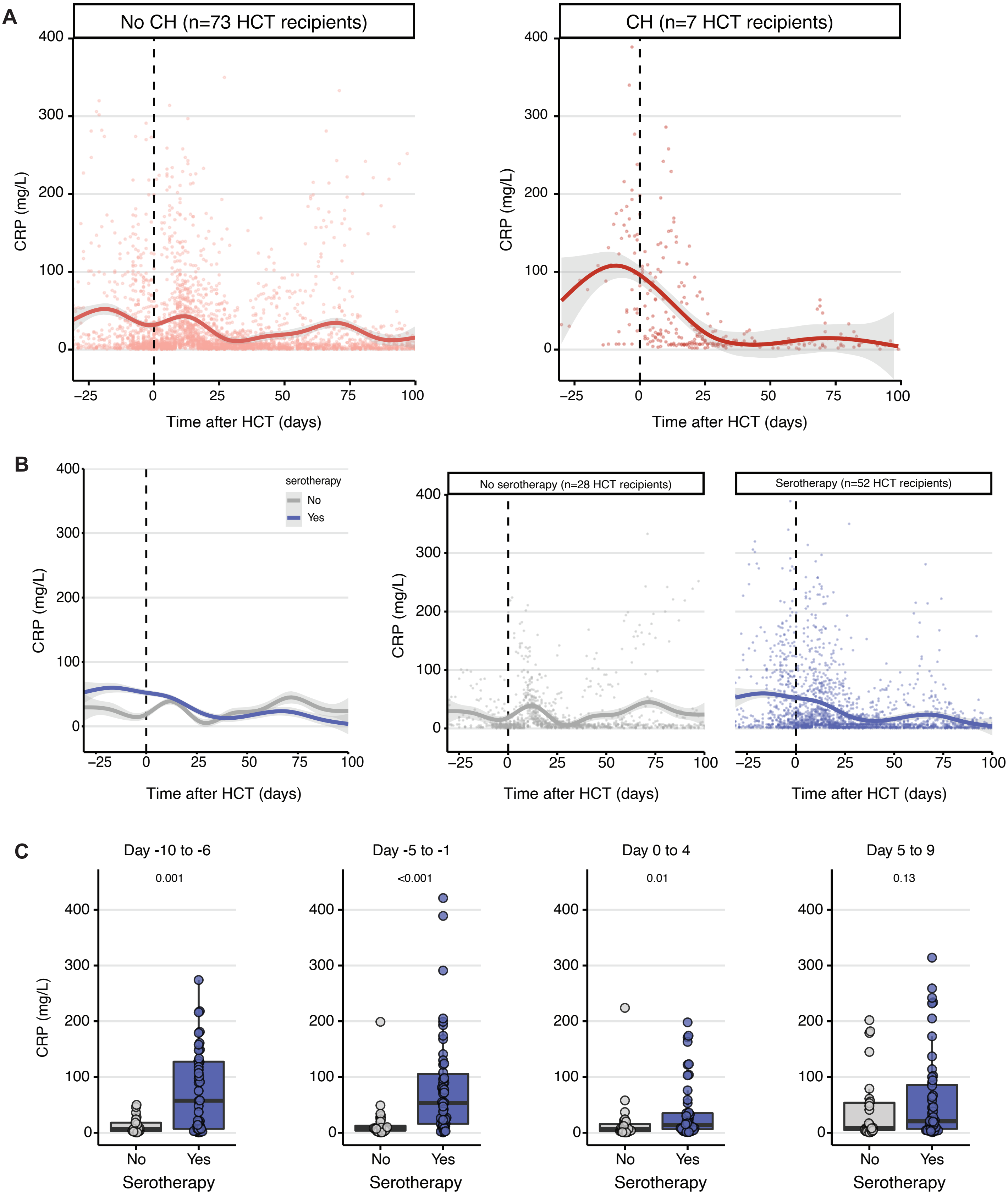


A) All CRP measurements around HCT in individuals with (left) and without CH (right). Lines indicate LOESS regression with 95% confidence interval
B) All CRP measurements around HCT in individuals that did (blue) or did not receieve serotherapy (grey). Lines indicate LOESS regression with 95% confidence interval.
C) Box plots demonstrating the highest CRP measured within 5-day intervals around graft infusion, split by serotherapy (yes/no). P values are calculated using Wilcoxon rank-sum test.

*Abbreviations: CH: clonal hematopoiesis; HCT: hematopoietic cell transplantation; CRP: C-reactive protein.*
